# Therapist-guided internet-delivered trauma-focused CBT for adolescents with PTSD: A feasibility trial

**DOI:** 10.64898/2026.01.18.26343775

**Authors:** Erica Mattelin, Hanna Weyler, Rebecca Andersson, Josefine Paulsen, Sandra Tielman, Anna Vikgren, Kristina Bondjers, Eva Serlachius, David Mataix-Cols, Maria Bragesjö

## Abstract

**Objectives:** Trauma-focused cognitive behavioural therapy (TF-CBT) is the established first-line treatment for paediatric posttraumatic stress disorder (PTSD), but access to evidence-based care remains limited. This study aimed to evaluate the feasibility and acceptability of a therapist-guided, 12-week, internet-delivered TF-CBT (iTF-CBT) programme for adolescents with PTSD, and to explore preliminary changes in PTSD symptoms.

**Design:** Single-group feasibility trial.

**Setting:** Save the Children, Sweden.

**Participants:** Twenty-two adolescents (13–17 years, 82% female) with primary PTSD.

**Interventions:** A 12-week, therapist-guided, internet-delivered TF-CBT comprising eight modules and parallel caregiver modules with joint child–caregiver activities.

**Outcomes:** Feasibility measures included recruitment pace, participant retention, treatment adherence (module completion), and therapist time. Acceptability was evaluated through satisfaction, credibility, negative effects, and reported adverse events. Preliminary treatment effects were evaluated as within-group changes in PTSD severity using independent evaluator-rated Clinician-Administered PTSD Scale (CAPS-CA-5) and the self-reported Child and Adolescent Trauma Screen 2 (CATS-2). Assessments occurred at baseline, during treatment, post-treatment, and at 1-month follow-up (primary endpoint).

**Results:** Recruitment was completed after seven months of active enrolment. Retention and adherence were high, satisfaction and credibility ratings were favourable, and no intervention-related serious adverse events occurred. Clinically meaningful within-group improvements were observed at the primary endpoint, with large reductions on CAPS-CA-5 (Cohen’s d = 1.27) and CATS-2 (Cohen’s d = 1.51).

**Conclusions:** Therapist-guided iTF-CBT for adolescents with PTSD was safe, feasible, acceptable, and associated with clinically meaningful symptom improvements. These findings support further evaluation in larger, controlled trials to determine efficacy, cost-effectiveness, and long-term outcomes.

**Trial registration:** ClinicalTrials.gov NCT06185244.

**Article Summary:** *Strengths and limitations of this study:*

- First internet-delivered TF-CBT trial for young people with PTSD
- Use of clinician-rated PTSD symptoms (CAPS-CA-5) in combination with validated self-report measures.
- The intervention was developed in close collaboration with clinicians, alongside contributions from young people.
- Absence of a control group.

## Introduction

Violence and other potentially traumatic events are common in adolescence; by late teens, nearly 60% of young people will have experienced at least one such event ^1^ ^2^. Among trauma-exposed children, the prevalence of post-traumatic stress disorder (PTSD) is estimated at 5–16% ^3^. In Sweden, where community shootings have become increasingly common ^4^, this proportion is expected to rise, as exposure to community violence is a well-established risk factor for PTSD ^5^.

PTSD is a debilitating condition characterised by intrusive memories, avoidance of trauma reminders, negative changes in cognition and mood, and hyperarousal, and is associated with marked functional impairment and psychiatric comorbidity ^6^. Untreated PTSD in youth substantially increases the risk of substance use, suicidality, poor mental health, and academic underachievement ^7–9^.

Trauma-focused cognitive behavioural therapy (TF-CBT) is a first-line treatment for paediatric PTSD ^10^, yet many adolescents do not receive treatment because of structural (limited clinician availability, waiting lists), logistical (travel, scheduling) and psychological (stigma, avoidance, caregiver burden) barriers. One possible solution to the limited availability of specialised TF-CBT for youth with PTSD is to deliver a low-intensity version of the treatment online with minimal remote support from a clinician. This kind of guided internet-delivered CBT differs from standard telepsychiatry in that the therapist does not actively deliver the treatment content in real time. Instead, the therapist provides minimal support asynchronously via a messaging system built in the online platform.

Adult trials suggest that therapist-guided internet-delivered CBT can achieve meaningful PTSD reductions and, in some settings, outcomes comparable to face-to-face care ^11–13^. While internet-delivered CBT is efficacious for several child and adolescent mental-health conditions ^14^ ^15^ therapist-guided internet-delivered TF-CBT (henceforth iTF-CBT) for adolescents with diagnosed PTSD remains understudied ^16^. Key uncertainties include feasibility (recruitment, retention, adherence), acceptability to adolescents and caregivers, therapist time and resource requirements, safety, and comparative evidence on efficacy and cost-effectiveness based on clinician- and self-reported outcomes.

The primary aim of this study was to evaluate the feasibility and acceptability of a therapist-guided, 12-week iTF-CBT programme for adolescents with PTSD. The main outcomes were recruitment pace, participant retention, treatment adherence, user satisfaction, and safety. The secondary aim was to estimate preliminary, within-group changes in both clinician-rated and self-reported PTSD symptom severity to inform outcome selection and power calculations for a future randomised controlled trial.

## Methods

### Study design and setting

This single-site, single group feasibility trial was delivered at a non-for-profit specialist clinic for children and young people (Save the Children Sweden). The clinic specialises in the assessment and treatment of vulnerable children exposed to violence, primarily presenting with child psychiatric problems such as PTSD. Referrals come from social services, schools and civil society organisations as well as through self-referral. Common to all cases is that the children have not accessed adequate care within the regular health care system. Independent evaluators administered the CAPS-CA-5 at baseline and at 1-month, the prespecified primary endpoint. We selected 1-month rather than immediate post-treatment because CAPS-CA-5 assesses the prior month, making post-treatment ratings largely overlap with treatment weeks; 1-month better captures stabilised change and permits timely transition to any further indicated care. The study was approved by the Swedish Ethical Review Authority (reference-number: 2023-06185-01), and registered at ClinicalTrials.gov before inclusion of the first participant (NCT06185244).

### Participants

Inclusion criteria were 13-17 years of age; primary diagnosis of PTSD; both children and parents fluent in Swedish; access to the Internet at home or ability to use study-provided prepaid mobile data vouchers; parent/guardian willing and able to take part in treatment. Exclusion criteria were psychosis or another primary psychiatric disorder of such severity requiring urgent specialist care or precluding participation (e.g., acute mania, an eating disorder requiring medical monitoring); ongoing substance abuse; high suicide risk; initiation or adjustment of any psychotropic medication within the last four weeks prior to treatment start and ongoing psychological treatment for PTSD.

### Sample size

This pilot was designed to evaluate feasibility and acceptability, not to establish efficacy. Consistent with guidance for feasibility studies, no formal power calculation was undertaken. A pragmatic target of N = 22 was chosen to enable estimation of key feasibility parameters and safety monitoring. Clinical outcomes were pre-specified as exploratory to provide preliminary effect size and variance estimates for a randomised trial.

### Primary outcome measures

#### Feasibility

Feasibility was assessed across four domains: study uptake, timeliness, data completeness, and resource use/cost. Study uptake comprised the recruitment rate (participants included per active week), screening-to-enrolment yield, and the proportion of eligible families who consented and initiated iTF-CBT and referral source (self-referred vs health care-signposted). Timeliness included the time between registration and the start of treatment. Data completeness was summarised at the 1-month primary endpoint. Resource use/cost quantified therapist workload, platform-logged outgoing messaging time per module and manually logged phone time and recruitment spend per enrolled participant.

#### Acceptability and satisfaction

Acceptability was indexed by module completion for adolescents and caregivers. Treatment discontinuation was defined a priori as completing fewer than four of 12 modules; conversely, completing ≥4/12 modules was taken to indicate receipt of the intervention’s core components. Acceptability was further measured by treatment satisfaction in both children and parents using the Client Satisfaction Questionnaire (CSQ) at post-treatment ^17^. The CSQ-8 consists of eight items, yielding a total score ranging from 8 to 32, with higher scores indicating greater satisfaction. The Credibility/Expectancy Questionnaire (CEQ) ^18^ was administered after the completion of treatment module 1. The questionnaire consists of six questions and range between 6-54, with higher scores indicating greater credibility.

#### Safety

Negative effects were measured with the Negative effects questionnaire (NEQ) by both adolescents and parents. NEQ ^19^ consists of 20 items with a total range of 0–80, with higher values representing more reported negative events. In addition to NEQ, therapists were encouraged to report other adverse events both at the weekly multidisciplinary team meetings but also directly to the research leader if they should occur during treatment. Safety was also monitored via a weekly platform-based suicidality item (“I have thoughts about death and about killing myself”) with predefined escalation (same-day clinician review, risk assessment, and referral), and by recording adverse events/serious adverse events (and whether or not they were judged to be treatment related) in the electronic health records.

### Secondary outcome measures

#### Clinician-rated outcomes

Changes in posttraumatic stress symptoms (primary measure of clinical efficacy) were assessed using the Clinician-Administered PTSD Scale for Children and Adolescents – Version 5 (CAPS-CA-5), the gold-standard clinical interview for diagnosing PTSD and evaluating symptom severity. Assessments were conducted at baseline and at 1-month follow-up. CAPS-CA-5 covers the 20 DSM-5 PTSD symptoms, associated distress/impairment, change since the prior assessment, and the dissociative specifier. Higher scores denote higher severity and impairment ^20^.

#### Self-report outcomes

All self-report secondary measures were administered via the digital platform. PTSD symptoms were assessed with the Child and Adolescent Trauma Screen – Version 2 ^21^, (CATS-2; range 0–60, higher scores representing greater symptom burden) at baseline, weekly during treatment, post-treatment, and at 1-month follow-ups. Depressive symptoms were assessed using the Mood and Feelings Questionnaire ^22^ (MFQ; 0–26, with higher scores indicating greater severity) at baseline, post-treatment, and at 1-month follow-ups. Functional impairment was evaluated with both youth- and parent-reported versions of the Work and Social Adjustment Scale ^23^, WSAS-Y and WSAS-P; 0–40, higher scores reflecting greater impairment) across the same time points. Health-related quality of life was measured with the KIDSCREEN-10 Index (10–50, higher scores denoting better quality of life) ^24^.

### Recruitment and procedures

Potential participants self-referred via social-media adverts linking to study information and a secure, password-protected study portal. Study staff then called registrants to provide further information, obtain consent to screening, and schedule an interview with a licensed psychologist (HW). Initial eligibility was checked against inclusion and exclusion criteria; non-eligible applicants were notified by phone and, where appropriate, referred to alternative health care services. Demographic information on adolescents (e.g., age, trauma type, gender, current and past psychotropic medication, and prior psychological treatment) was also collected, while parental data were obtained separately through an online questionnaire. Eligible dyads completed baseline questionnaires online including CATS-2, MFQ, WSAS-Y/P, TIC-P, KIDSCREEN-10 prior to clinical assessment. Diagnostic assessment comprised the Mini International Neuropsychiatric Interview for Children and Adolescents (MINI-KID) ^25^ to rule out alternative primary diagnoses and identify comorbidities, and the CAPS-CA-5 to confirm PTSD and rate past-month severity. Interviews were conducted in person or via video/telephone by a clinical psychologist or trained psychology student. Baseline assessments additionally included suicide risk (MINI-KID) and global functioning with The Children’s Global Assessment Scale (CGAS).

Final inclusion was confirmed at weekly multidisciplinary case conferences. Treatment typically commenced the following week; some start dates were deferred over the summer to avoid interruption. Both clinician-rated and self-reported outcomes were assessed at baseline and 1-months assessment. Written informed consent was obtained from adolescents and their legal guardians. Non-eligible applicants were notified by phone and, if needed, referred to regular health care.

### Intervention

The iTF-CBT protocol mirrors first-line treatment for children with PTSD ^26^ and is delivered via a secure digital platform. The programme comprises eight adolescent modules with parallel caregiver modules (Table 1). Core components include psychoeducation, affect/emotion regulation, trauma narration and cognitive processing, in-vivo exposure, safety planning, and relapse prevention. Treatment initially targets the most distressing trauma memory, with flexibility to address additional events as needed. To maintain continuity in graded sharing (i.e., progressive disclosure of the trauma narration from outline to fuller detail as tolerated), we scheduled the joint caregiver–adolescent session prior to initiating vivo exposure.

**Table 1.** Overview of the treatment content^1^.

| **Week** | **Child** | **Parent** | **Joint** |
| --- | --- | --- | --- |
| 1 | Psychoeducation<br>Parents and children learn about exposure to trauma and the potential effects | Parental skills and psychoeducation | Psycheducation |
| 2 | Relaxation<br>Relaxation techniques are taught to help manage stress and anxiety and to prepare for the trauma narration | Parental skills and relaxation | Relaxation |
| 3 | Affective modulation<br>Involves identifying, labeling, expressing and coping with feelings. | Parental skills and affective modulation | Affective regulation |
| 4 | Cognitive coping<br>Involves how thoughts, feelings and behaviours are connected and how we ourselves can influence these. | Parental skills and cognitive coping | Cognitive coping |
| 5 | Trauma narration and cognitive processing<br>Revisiting trauma memory through narrative exposure and cognitive processing. | Parental skills and cognitive processing | Cognitive processing |
| 6 | Trauma narration and cognitive processing | Parental skills and cognitive processing | Cognitive processing |
| 7 | Trauma narration and cognitive processing | Parental skills and cognitive processing | Cognitive processing |
| 8 | Trauma narration and cognitive processing | Parental skills and cognitive processing | Cognitive processing |
| 9 | Joint child-parent session<br>Joint session in which the child shares the whole trauma narrative | Parental skills and cognitive processing | Cognitive processing |
| 10 | In vivo<br>Gradually confronting and overcoming fears and reminders of the trauma through real-life exposures | In vivo | In vivo |
| 11 | Enhancing safety and future development<br>Safety planning and maintenance planning | Future safety | Future safety |
| 12 | Future safety | Future safety | Celebration |

<sup>1</sup>All modules involve gradual exposure.

Modules use age-appropriate text, audio, and brief video vignettes and are accessed sequentially (one module per week) with home practice tasks. Each adolescent and their caregiver/guardian were assigned a named therapist who provided guidance primarily via secure asynchronous messaging on the platform. Caregivers engage in parallel modules that mirror the child’s content, including parenting practices (such as praise and validation), with joint caregiver–child activities integrated throughout. The intervention was delivered in Swedish.

### Therapists

Seven clinical psychologists/psychotherapists from Save the Children delivered the intervention. All had prior TF-CBT training (3 to 12 years of experience with the method) and received onboarding to the digital protocol and platform before treatment started. Therapists were expected to log in at least twice per week. The therapists attended weekly joint case conferences with the Principal Investigator (PI; MB) to secure treatment fidelity, monitor safety for the participants and identify adjustment needs.

### Assessors

Outcome assessments were conducted primarily by psychology students in the final semester of a five-year master’s programme, all trained in CGAS, MINI-KID, and CAPS-CA-5 prior to data collection. In addition, three therapists conducted a subset of assessments after completing the same training. Given the non-randomised design, blinding of outcome assessors was not feasible.

### Patient and public involvement

Stakeholder collaboration spanned all stages and included adolescents with lived experience of PTSD who reviewed the intervention and provided iterative feedback on content, language, and usability, alongside child and adolescent psychiatry personnel and Save the Children clinicians. Needs were jointly identified and prioritised by Save the Children staff and health care professionals, highlighting gaps in effective treatment and barriers to access.

### Statistical methods

Feasibility and acceptability metrics were summarised descriptively. For the clinical outcomes, Cohen’s *d* statistics were derived from linear mixed-effect models with random intercepts for participants. Differences between time points were based on estimated marginal means (EMMs), and effect sizes were standardised using the pre-treatment standard deviation. All analyses were conducted in R 4.5.1.

### Ethics and consent

This study was approved by the Swedish Ethical Review Authority (2023-06185-01). All participants and their legal guardian/s provided written consent before participation.

## Results

### Feasibility

#### Study uptake

Between March and December 2024, 158 families registered interest (Table 3). Recruitment was paused during June–August, yielding seven months of active recruitment. Approximately 23% of enquiries were prompted by a health care-provider recommendation to self-refer to the study. During active months, the mean recruitment rate was 0.8 participants per week.

Of the 158 registrants, 85 adolescents/caregivers (54%) completed a telephone screen, and 36 (42% of those screened) completed the CAPS-CA-5 assessment; 22 adolescents from across Sweden were enrolled (61% of those assessed). Baseline demographic and clinical characteristics are summarised in Table 2, and Figure 1 shows participant flow. There were no study drop-outs.

**Figure 1.**
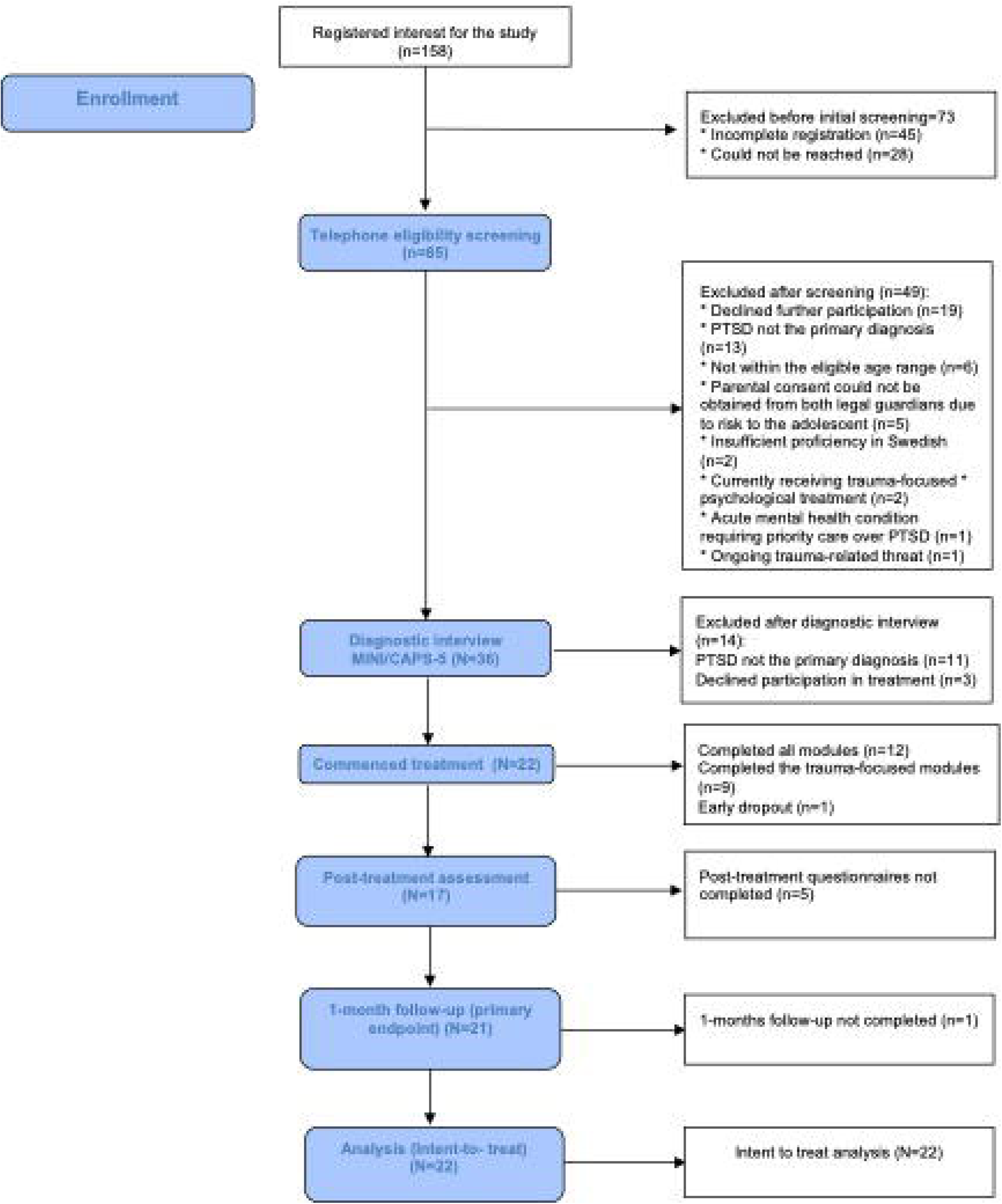
CONSORT flow diagram of participant progress through the study. MINI-KID = Mini International Neuropsychiatric Interview for Children and Adolescents; CAPS-CA-5 = Clinician-Administered PTSD Scale for Children and Adolescents for DSM-5.

**Table 2.** Baseline demographic and clinical characteristics of the sample.

|  | Total |
| --- | --- |
| **Age, mean. (SD), min-max** | 16.06 (1.52), 13-17 |
| **Gender n (%)** |  |
| Male | 3 (13.6) |
| Female | 18 (81.8) |
| Non-binary | 1 (4.5) |
| **Main contact person, mothers, n (%)** | 21 (95.5) |
| **Education of primary contact person n (%)** |  |
| Secondary education | 4 (13.6) |
| Post-secondary education (not tertiary/college) | 3 (13.6) |
| Higher education (3 years or less) | 4 (18.2) |
| Higher education (3<) | 11 (50.0) |
| **Current psychiatric comorbidity n (%)** |  |
| At least one psychiatric comorbidity<sup>1</sup> | 22 (100) |
| ADHD/ASD | 15 (68.2) |
| **Type of trauma n (%)** |  |
| Sexual violence | 5 (22.7) |
| Witnessed a medical emergency | 2 (9.1) |
| Physical assault | 4 (18.2) |
| Witnessed violence | 3 (13.6) |
| Bullying | 4 (18.2) |
| Other | 4 (18.2) |
| **Number of reported traumatic events, mean (SD)** | 4.36 (2.7) |
| **Current use of any psychotropic medication n (%)** | 13 (59.1) |
| **Suicidality<sup>1</sup> n (%)** |  |
| Low risk | 14 (63.6) |
| Medium risk | 3 (13.6) |
| High risk | 5 (22.7) |

<sup>1</sup> Comorbidity and suicidality was assessed using the Mini International Neuropsychiatric Interview for Children and Adolescents (MINI-KID 7.0). <sup>2</sup> ADHD/ASD refers to screen-positive results for attention-deficit/hyperactivity disorder or autism spectrum disorder in MINI-KID. PTSD = post-traumatic stress disorder.

**Table 3.** Feasibility and acceptability outcomes.

| Domain | Indicator | Result | Comment |
| --- | --- | --- | --- |
| Recruitment period | Active accrual months | March–December 2024 (paused June–August) | Seven months of active recruitment |
| Registrants | Families registered interest | 158 | Nationwide sample |
| Screening | Completed telephone screen | 85 (54 % of registrants) |  |
|  | Completed CAPS-CA-5 assessment | 36 (42 % of those screened) |  |
| Enrolment | Enrolled participants | 22 (61 % of those assessed) | No study drop-outs |
| Accrual rate | Mean enrolment per week | 0.8 participants per week | Active months only |
| Therapists | Number of therapists delivering iTF-CBT | 7 | Each treated 2–4 cases |
| Treatment duration | Planned treatment period | 12 weeks | Minor deviations ($\pm 1$–2 weeks due to illness) |
| Timeliness | Mean time registration → treatment | 95.7 days (50.7 excluding summer delay) |  |
| Data completeness | 1-month follow-up | 21 / 22 ($\approx 95$ %) completed | Final assessment 6 May 2025 |
| Resource use | Mean asynchronous messaging time per module | Adolescents = 8.90 min (SD 0.25); Caregivers = 9.44 min (SD 3.46) |  |
|  | Mean phone/video | Adolescents = 1.91 | 10 adolescents and |
| | calls per recipient | (SD 3.07); Parents = 1.36 (SD 1.83) | 13 caregivers had ≥ 1 call |
| Recruitment cost | Estimated per participant | ≈ 151 GBP | |
| Adherence | Modules completed (mean of 12) | Adolescents = 10.5 (SD 2.33); Caregivers = 10.5 (SD 2.48) | 50 % adolescents and 54.5 % caregivers completed all modules |
| Acceptability | Discontinued treatment | 1 adolescent–caregiver dyad (4.5 %) |  |
| Credibility / expectancy | Mean (SD) | Adolescents = 34.32 (6.70); Parents = 41.05 (8.62) | Measured using the Credibility/Expectancy Questionnaire (CEQ) |
| Satisfaction (CSQ-8) | Mean (SD) | Adolescents (n=17) = 24.18 (5.13); Parents (n=20) = 26.95 (3.25) | Item-level means 2.41–3.53 (adolescents); 2.80–3.85 (parents) |

Values are presented as mean (SD) unless otherwise stated. iTF-CBT = internet-based trauma-focused cognitive behavioural therapy; CAPS-CA-5 = Clinician-Administered PTSD Scale for Children and Adolescents for DSM-5; PTSD = post-traumatic stress disorder; CSQ = Client Satisfaction Questionnaire; CEQ = Credibility/Expectancy Questionnaire.

#### Timeliness

The mean time from registration to treatment was 95.7 days. However, a substantial portion of participants experienced a delay due to the Swedish summer break. When excluding this group, the mean registration to treatment time decreased to 50.7 days.

#### Data completeness

At the 1-month follow-up primary endpoint, outcome data were available for 21/22 participants (≈ 95% completeness). The final 1-month assessment was completed on 6 May 2025.

#### Resource use and cost

Average asynchronous messaging time per module was 8.90 min for adolescents (SD = 0.25) and 9.44 min (SD = 3.46) min for caregivers. Ten adolescents and thirteen caregivers had ≥1 phone or video call; the mean number of calls was 1.91 (SD = 3.07) for adolescents and 1.36 for parents (SD = 1.83). Recruitment costs were estimated at approximately 151 GBP per participant.

### Acceptability of iTF-CBT

#### Treatment adherence

On average, 10.50 of 12 modules were completed by both adolescents (SD = 2.33) and caregivers (SD = 2.48). By the end of treatment, 11 adolescents (50%) and 12 caregivers (55%) completed all modules. One adolescent–caregiver dyad discontinued treatment.

#### Credibility and satisfaction

Average treatment credibility/expectancy was 34.32 (SD = 6.70) for adolescents and 41.05 (SD = 8.62) for parents. Mean treatment satisfaction for adolescents was 24.18 (SD = 5.13), indicating a generally high level of satisfaction with the intervention. The corresponding numbers for parents were 26.95 (SD = 3.25). Item-level means ranged from 2.41 to 3.53 (adolescents) and 2.80 to 3.85 (parents) on the 4-point scale, suggesting consistently positive experiences across most satisfaction domains.

#### Negative effects and adverse events

Between baseline and the 1-month follow-up, a total of 196 negative treatment-related effects were reported by both adolescents and parents (Table 4). Adolescents reported 102 treatment-related events, with a mean impact rating of 1.91 on a 0–4 scale. The most reported event was unpleasant thoughts. Parents reported 89 events, with a mean impact rating of 2.12. The most frequent events reported by parents were lack of motivation for treatment and unpleasant thoughts.

**Table 4.** Negative effects and adverse events.

| Category | Definition / example | Adolescents n (%) | Parents n (%) |
| --- | --- | --- | --- |
| Negative effects (NEQ) | Total treatment-related effects reported between baseline and 1-month follow-up (NEQ completers: adolescents n = 17) and parents = 20) | 102 events (treatment-related). Mean impact 1.91. | 89 events (treatment-related). Mean impact 2.12 |
| Most common NEQ items | Unpleasant thoughts | 16 (94.1%) | 13 (64.7%) |
|  | Lack of motivation for treatment | 11 (55.0) | 13 (65.0) |
| Adverse events (weekly self-report) | ≥ 1 adverse event during treatment (open-ended AE item) | 9 / 22 (40.9 %) | 10 / 22 (45.5 %) |
| Most frequent AEs | Increased anxiety / tiredness / feeling low or depressed | 9.1 % | 22.7 % anxiety, 13.6 % low mood |
| Suicidal ideation | Item ≥ 3 (“I have thoughts about death and about killing myself”) | 1 (4.5 %) | — |
| Serious adverse event (SAE) | Hospital admission | 1 (4.5 %) | — |

Table note: Values are numbers (%) unless otherwise stated. Mean impact scores range 0 (“no impact”) to 4 (“very severe impact”). NEQ = Negative Effects Questionnaire; AE = adverse event; SAE = serious adverse event.

Based on the weekly open-ended adverse events question, 40.1% of adolescents reported at least one adverse event (median=0, range 0-7) during treatment. The most common were increased anxiety, fatigue, and feeling low or more depressed (9%). Among parents, 45% reported at least one adverse event, most frequently increased anxiety and worry (23%) or feeling low (14%). Suicidal ideation (score ≥3 on the item: “I have thoughts about death and about killing myself”) was reported by one adolescent. A single serious adverse event (SAE) occurred (hospital admission), that investigators judged unrelated to the intervention.

### Clinical outcomes

#### Preliminary outcome measures

A linear mixed-effects model showed a significant change in symptoms of PTSD (CAPS-CA) between baseline and 1-month follow-up (Table 5, B = -12.04, p <0.001, 95% CI [-16.2 to -7.83], p < 0.001). Within-group Cohen’s *d* was 1.27 (95% CI [0.85 to 1.68]).

**Table 5.** Mean scores (SD) on clinician- and self-reported outcome measures at baseline, post-treatment, and 1-month follow-up.

| Measure | M (SD) Range |
| --- | --- |
| **Clinician rated** |  |
| CAPS-CA-5 |  |
| Baseline (N=22) | 34.68 (SD = 9.51), 19–52 |
| 1-month post (N=21) | 23.05 (SD = 10.51), 5–40 |
| **Self- and parental assessed** |  |
| CATS-2 Child |  |
| Baseline (N=22) | 36.64 (SD = 4.69), 26–44 |
| Post-treatment (N=17) | 28.76 (SD = 11.76), 0–42 |
| 1-month post (N=17) | 29.71 (SD = 10.62), 8–47 |
| CATS-2 Parent |  |
| Baseline (N=22) | 32.41 (5.18) 24-47 |
| Post-treatment (N=20) | 20.90 (11.39) 0 - 50 |
| 1-month post (N=20) | 19.05 (11.48) 0-42 |

Values are presented as mean (SD). Higher scores indicate greater symptom severity. CAPS-CA = Clinician-Administered PTSD Scale for Children and Adolescents; CATS-2-C = Child and Adolescent Trauma Screen (child report); CATS-2-P = Child and Adolescent Trauma Screen (parent report).

#### Secondary treatment outcomes

Linear mixed models showed a decrease in self-reported symptoms of post-traumatic stress (CATS-2-C) between baseline and 1-month follow-up (Table 5, B = −7.09, *p* < .001, 95% CI [−12.23 to −1.94]). The within-group effect size was large (Cohen’s *d* = 1.51 95% CI [0.41 to 2.61]). For parental reports of post-traumatic stress symptoms (CATS-2-P), there was likewise a large and statistically significant reduction (B = −13.21, *p* < .001, 95% CI [−17.83 to −8.59]), corresponding to a very large within-group effect (Cohen’s *d* = 2.55, 95% CI [1.66 to 3.44]).

For child-reported well-being (KIDSCREEN-C), there was a large and statistically significant improvement (Table S1, B = 10.67, *p* < .001, 95% CI [7.85 to 13.50]); the within-group effect size was very large (Cohen’s *d* = 2.44, 95% CI [1.80 to 3.09]). In contrast, for parental reports of child well-being (KIDSCREEN-P), there was no significant change (B = 0.77, *p* = .38, 95% CI [−0.97 to 2.51]); the corresponding effect size was small (Cohen’s *d* = 0.17, 95% CI [−0.21 to 0.54]).

For child-reported functional impairment (WSAS-Y), there was no significant change from baseline to 1-month follow-up (B = 1.73, *p* = .29, 95% CI [−1.56 to 5.02]); the within-group effect size was small (Cohen’s *d* = 0.22, 95% CI [−0.20 to 1.64]). For parental reports of child functioning (WSAS-P), there was a moderate and statistically significant improvement (B = -4.71, *p* = .006, 95% CI [-8.02 to -1.40]), corresponding to a moderate within-group effect (Cohen’s *d* = 0.50, 95% CI [0.15 to 0.86]).

For child-reported depressive symptoms (MFQ-C), there was no significant change (B = −0.52, *p* = 0.69, 95% CI [−2.16 to 3.21]); the effect size was small (Cohen’s *d* = −0.13, 95% CI [−0.54 to 0.80]). For parental reports of depressive symptoms in their children (MFQ-P), there was a large and statistically significant reduction (B = −3.36, *p* < .001, 95% CI [−5.17 to −1.56]); the within-group effect size was large (Cohen’s *d* = 0.88, 95% CI [0.41 to 1.36]).

## Discussion

This pilot study aimed to evaluate the feasibility, acceptability, safety, and preliminary clinical effects of a therapist-guided, internet-delivered TF-CBT (iTF-CBT) programme for adolescents with diagnosed PTSD, delivered within a non-for-profit specialist clinic for children and young people operated by Save the Children Sweden, with parallel caregiver access. Overall, the intervention was feasible, well accepted, and associated with promising within-group improvements.

Retention and adherence were strong; credibility and satisfaction ratings were favourable; and no intervention-related serious adverse events were observed. These findings align with prior evidence for internet-delivered interventions in adult PTSD ^11^ ^12^ and extend the paediatric iCBT literature by targeting adolescents with diagnosed PTSD.

Recruitment primarily relied on advertising directed at parents. While this approach was adequate, it may have been faster and more effective if it had involved direct engagement with youth. Future trials should explore youth-centred outreach strategies such as advertisements directed to the youth instead of parents and referrals through health care and other services. Despite these barriers, inclusion rates among assessed youth were high.

High treatment adherence and regular therapist contact indicate that asynchronous, message-based guidance with optional calls is feasible for both families and clinicians. All participants screened for at least one additional diagnosis, and about two-thirds screened positive for ADHD or ASD. Despite this high comorbidity and associated dropout risk, over 95% maintained regular therapist contact across 12 weeks, demonstrating strong engagement. As expected in exposure-based care, short-term increases in distress were common but typically transient and treatment-concordant. One serious adverse event occurred but was deemed unrelated to treatment. This underscores the importance of ongoing risk monitoring and predefined escalation pathways in digital PTSD treatment.

The digital format was generally well received by both adolescents and caregivers. Average therapist time was approximately 18 minutes per family per week, substantially lower than the 90 minutes typically required for face-to-face TF-CBT and similar to earlier iCBT studies ^27^. If confirmed in future trials, this efficiency gain would have clear clinical and health-economic relevance.

Within-group analyses showed large clinician-rated improvements and moderate adolescent self-reported improvements, whereas parents reported larger reductions across several domains. Such discrepancies between informants are common in the literature on PTSD but also other conditions ^28^ ^29^. They underscore the importance of using multiple informants when evaluating treatment effects in youth PTSD. One participant showed symptom worsening (>5 points on CAPS-CA-5) and required stepped-up care. This underscores the need for ongoing monitoring and timely referral to more intensive treatment in future trials.

This study has several strengths, including the use of CAPS-CA-5 as the gold-standard outcome, robust safety monitoring, and delivery in a non-profit specialist care clinic setting with high retention. Limitations include the lack of a control group, self-selection of participants, and provision in Swedish only, limiting generalisability to migrant and refugee populations. Further, improved recruitment pace will be necessary in a future RCT.

## Conclusions

Therapist-guided internet-delivered TF-CBT for adolescents with PTSD is feasible, acceptable, safe, and associated with promising clinical outcomes in a routine care setting. Future randomised controlled trials are warranted to establish its clinical efficacy, cost-effectiveness, and long-term outcomes.

## Data Availability

The datasets generated and analysed during the current study are not publicly available due to Swedish and EU data protection regulations but are available from the corresponding author on reasonable request, provided that data sharing is compliant with applicable legal requirements.

## Declarations / Additional files

## Acknowledgements

We are grateful to the participating children and parents for their involvement in the study. We also acknowledge the children, caregivers, and therapists who contributed to the development of the treatment. In addition, we acknowledge the contributions of Martin Herrgård, Sofia Michael, Laura Korhonen (Director of Barnafrid), Sofia Bidö (Director of Save the Children’s Centre for Support and Treatment), and Jannes Grudin.

## Author statement

Conceptualization: EM, ES, DMC, MB

Methodology: EM, ES, DMC, MB, KB

Intervention development: EM, ST, HW, JP, AV, MB

Formal analysis / analysis plan: EM, RA

Investigation / Data collection: EM, ST, HW, JP, AV

Writing – original draft: EM

Writing – review & editing: EM, ES, DMC, MB, ST, HW, JP, AV, KB, RA

Supervision: MB, DMC

