## Supplementary Table 1 for "Therapist-guided internet-delivered trauma-focused CBT for adolescents with PTSD: A feasibility trial"

Table S1. Means (SD) for all measures at the three assessment points

| Measure | M (SD) |
| --- | --- |
| **Self- and parental assessed** |  |
| KIDSCREEN-10 Child |  |
| Baseline | 19.45 (4.54) 13-28 |
| Post-treatment | 20.24 (5.13) 11-28 |
| 1-month post | 31.24 (6.69) 21-44 |
| KIDSCREEN-10 Parent |  |
| Baseline | 20.14 (4.65) 13-30 |
| Post-treatment | 20.80 (4.74) 14-32 |
| 1-month post | 21.20 (5.79) 11-31 |
| WSAS Child |  |
| Baseline (N=21) | 17.71 (7.87) 4-30 |
| Post-treatment (N=16) | 20.12 (9.19) 3-30 |
| 1-month post (N=16) | 19.00 (9.85) 1-35 |
| WSAS Parent |  |
| Baseline(N=21) | 18.24 (9.37) 3-36 |
| Post-treatment(N=19) | 14.53 (10.92) 0-35 |
| 1-month post(N=19) | 13.63 (11.48) 0-36 |
| MFQ Child |  |
| Baseline (N=22) | 12.36 (4.01) 6-22 |
| Post-treatment(N=17) | 10.94 (5.27) 1-19 |
| 1-month post (N=17) | 11.76 (6.55) 0-24 |
| MFQ Parent |  |
| Baseline (N=22) | 9.41 (3.81) 1-15 |
| Post-treatment(N=20) | 7.05 (4.91) 0-17 |
| 1-month post (N=20) | 5.90 (4.17) 0-14 |

Values are presented as mean (SD). Higher scores on the KIDSCREEN indicate greater well-being, whereas higher scores on the MFQ and EWSAS indicate greater symptom severity and functional impairment. KIDSCREEN-C = KIDSCREEN, child report; KIDSCREEN-P = KIDSCREEN, parent report; MFQ-C = Mood and Feelings Questionnaire, child report; MFQ-P = Mood and Feelings Questionnaire, parent report; EWSAS-C = Everyday Social and Academic Stress Scale, child report; EWSAS-P = Everyday Social and Academic Stress Scale, parent report.
